# Non-alcoholic steatohepatitis patients exhibit reduced CD47, and increased sphingosine, cholesterol and MCP1 levels in the erythrocyte membranes

**DOI:** 10.1101/2022.01.12.22269197

**Authors:** Charalampos Papadopoulos, Eleftheria Spourita, Konstantinos Mimidis, George Kolios, Ioannis Tentes, Konstantinos Anagnostopoulos

## Abstract

Non-alcoholic steatohepatitis (NASH) constitutes a significant cause of deaths, liver transplantations and economic costs worldwide. Despite extended research, investigations on the role of erythrocytes are scarce. Red blood cells from experimental animals and human patients with NASH, present phosphatidylserine exposure which is then recognized by Kupffer cells. This event leads to erythrophagocytosis, and amplification of inflammation through iron disposition. In addition, it has been shown that erythrocytes from NASH patients release the chemokine MCP1, leading to increased TNF-α release from macrophages RAW 264.7. However, erythrophagocytosis can also be caused by reduced CD47 levels. In addition, increased MCP1 release could be either signal-induced, or caused by higher MCP1 levels on the erythrocyte membrane. Finally, erythrocyte efferocytosis could provide additional inflammatory metabolites. In this study, we measured the erythrocyte membrane levels of CD47 and MCP1 by ELISA, and cholesterol and sphingosine with thin-layer chromatography. 18 patients (8 men, 10 women aged 56.7±11.5 years) and 14 healthy controls (7 men, 7 women aged 39.3±15.5 years) participated in our study. The erythrocyte CD47 levels were decreased in the erythrocyte membranes of NASH patients (844±409 pg/ml) compared to healthy controls (2969±1936 pg/ml) with P(Healthy>NAFLD)=99.1%, while the levels of MCP1 were increased in NASH patients (389±255 pg/ml), compared to healthy controls (230±117 pg/ml) with P(Healthy<NAFLD)=88.9%. Moreover, in erythrocyte membranes there was a statistically significant accumulation of sphingosine and cholesterol in NASH patients, compared to healthy controls. Our results imply that erythrocytes release chemotactic “find me” signals (MCP1), while containing reduced “do not eat me” signals (CD47). These molecules can lead to erythrophagocytosis. Next, increased “goodbye” signals (sphingosine and cholesterol) could augment inflammation by metabolic reprogramming.

## Introduction

The global financial growth has led to a substantial increase of sedentary life^1^. As such, there is a remarkable rise in obesity and metabolic syndrome^2^. Nonalcoholic steatohepatitis (NASH), a frequently occurring hepatic manifestation of metabolic syndrome^3^ constitutes a significant cause of deaths, liver transplantations^4^ and economic cost^5^, worldwide. Despite extended research on the molecular pathogenesis, there is still need for the discovery of therapeutic targets. So far, many cell types have been investigated for their contribution to the pathogenesis of nonalcoholic fatty liver disease and its more severe inflammatory form, NASH. The role of platelets^6^, monocytes/macrophages, hepatocytes, hepatic stellate cells, endothelial cells, neutrophils, T lymphocytes, natural killer cells and other innate lymphoid cells has been extensively studied^7,8^. However, research on the role of erythrocytes has remained scarce.

Erythrocytes are important modulators of the adaptive and innate immunity, and participate in the systemic metabolism of lipids^9^. In addition, erythrocyte function is heavily influenced by toxic lipid mediators^10^, hormones^11^ and molecular constituents of inflammation, all important hallmarks of NASH pathogenesis^12^. Otogawa et al^13^, have shown that erythrocytes from both experimental animals and human patients with NASH, accumulate in the liver. This is caused by oxidative stress-induced phosphatidylserine exposure, which is then recognized by Kupffer cells. This event leads to erythrophagocytosis, amplifying inflammation. It was recently shown that erythrocytes from NASH patients release the chemokine MCP1, without significant changes in the release of other cytokines and of the signaling lipids, sphingosine 1 phosphate (S1P) and lysophosphatidic acid (LPA). This event was followed by increased TNF-α release from macrophages RAW 264.7^14^.

Erythrophagocytosis can also be caused by reduced CD47 levels^15^. In addition, increased MCP1 release could be either signal-induced, or caused by higher MCP1 levels on the erythrocyte membrane. Finally, erythrocyte efferocytosis could provide inflammatory metabolites^16^. Hence, in this study, we tried to elaborate on these mechanisms by measuring the levels of CD47, MCP1, cholesterol and sphingosine on the erythrocyte membrane of NASH patients and healthy controls.

## Methods

### Patients and healthy controls

18 patients (8 men, 10 women) and 14 healthy controls (7 men, 7 women) participated in the study. They were recruited by the 1st Pathology Clinic, Department of Medicine, Democritus University of Thrace, Alexandroupolis, Greece. All patients presented hepatic steatosis according to ultrasonography. After exclusion of viral, alcoholic, drug and other causes, patients were evaluated by non-invasive biomarkers (NFS, FIB4, AST/ALT ratio). Their anthropometric and clinical characteristics are shown in Table 1. The study was approved by the Scientific Council of the University Hospital of Alexandoupolis and the Ethics Committee, after informed consent of the participants.

**Table 1.** Anthropometric and clinical characteristics of NASH patients and healthy subjects. P(Healthy<NAFLD) or P(Healthy>NAFLD) is the probability that the difference between means of healthy subjects and NAFLD patients if less or greater than zero, respectively.

| Parameter | Healthy controls (n=14) | NASH patients (n=18) | P(Healthy<NASH) or P(Healthy>NASH) |
| --- | --- | --- | --- |
| Age (years) | 39.3±15.6 | 56.7±11.5 | 97.9% |
| BMI<sup>a</sup> | 25.3±3.8 | 32.9±4.3 | 99.9% |
| AST (U/L) | 22±9 | 48±19 | 98.0% |
| ALT (U/L) | 22±7 | 82±68 | 95.0% |
| Triglycerides (mg/dL) | 152±18 | 224±160 | 96.3% |
| Cholesterol (mg/dL) | 154±14 | 197±47 | 92.7% |
| HDL-Cholesterol (mg/dL) | 65±14 | 45±7 | 99.5%<sup>c</sup> |
| LDL-Cholesterol (mg/dL) | 59±8 | 110±43 | 97.6% |
| Platelets (thousands/μl) | 240±54 | 223±63 | 52.8%<sup>c</sup> |
| FIB4<sup>b</sup> | 0.77±0.35 | 1.90±1.28 | 97.2% |
| AST/ALT | 0.97±0.16 | 0.91±0.50 | 32.0%<sup>c</sup> |

<sup>a</sup> Body Mass Index, <sup>b</sup> FIB4: Non-Alcoholic Fatty Liver Disease Fibrosis score
<sup>c</sup> Most probabilities are for P(Healthy<NAFLD). The probabilities marked by <sup>c</sup> are P(Healthy>NAFLD).

### Erythrocyte isolation

Three milliliters of blood containing 5.4 mg EDTA was centrifuged at 200×g for 10 minutes at 4°C. The plasma and buffy coat were removed. Then, 1 milliliter of erythrocyte pellet was washed with cold saline solution and centrifuged at 200×g for 10 minutes at 4°C. This step was repeated 4 times, and red blood cells were then collected from the bottom of the tube.

### Erythrocyte membrane isolation

Erythrocytes were diluted 1:10 (v/v) with cold hemolysis solution (Tris 1 mM–NaCl 10 mM – EDTA 1 mM pH 7.2) followed by incubation for 30 minutes at 4°C with continuous shaking. They were then centrifuged at 15000 rpm in a HERMLE 323 K centrifuge, 220.80 VO2 rotor, at 4°C for 15 minutes. The pellet was collected, and the last step was repeated as many times as needed, until the final pellet was milky-white, signifying hemoglobin removal. Samples were kept at −80°C until analysis.

### Production of erythrocyte-conditioned media

Erythrocytes (5×10^7^/ml) were incubated in RPMI 1640, containing 10% FBS (v/v), 1% streptomycin/penicillin (v/v), at 5% CO2, 37°C, for 24 hours. Then, the erythrocyte-derived conditioned medium (ECM), from both patients (P-ECM) and healthy controls (H-ECM) was collected with centrifugation at 200×g for 10 minutes. Conditioned media were stored at −80°C. As control, growth medium was placed in 6 well plates, in the same conditions, thereafter, following the same procedures as for ECM. Spectrophotometric assay indicated that no hemoglobin was released by erythrocytes, excluding thus hemolysis.

### Inhibition of sphingosine kinase

Erythrocytes (10^9^/ml) were incubated in RPMI 1640, containing 10% FBS (v/v), 1% streptomycin/penicillin (v/v), at 5% CO2, 37°C, for 24 hours, with or without N,N-dimethylsphingosine (DMS), which is an inhibitor of sphingosine kinase, at a concentration of 14 μM.

### Lipid extraction

For lipid extraction, 500 μL of ECM or erythrocyte membrane was used. We used a modified Folch method^17^. Briefly, 500μl of 2:1 chloroform/methanol were added to 500μl of ECM or erythrocyte membrane. Then, the sample was vortexed for 30 seconds, and centrifuged for 10 minutes at 5000×g. The organic phase was collected, and the previous step was repeated to the aqueous phase. The two organic phases were washed with water by centrifugation for 10 minutes at 5000×g, and then evaporated at 70° C.

### Lipid separation and visualization

Thin layer chromatographic analysis of lipids was done on a 10×10 cm chromatographic plate (TLC Silica gel 60 F254 (Merck KGaA 64071 Darmstadt, Germany) using a mixture of chloroform/methanol/acetic acid/ water (60/50/1/4) (v/v/v/v). Before loading the samples, the plate was desiccated at 150° C for 10 minutes and pre-run with the developing solvent mixture. After sample separation, the plate was dried with hot air and then placed in a closed container of vaporized iodine (3.5 gr), placed facing the bottom of the container, for 30 minutes at room temperature. Lipids appeared as dark yellow bands against a lighter background. In order to ensure that the various lipids were well separated, they were identified by comparison with lipid standards, namely cholesterol, phosphatidylethanolamine, phosphatidylinositol, phosphatidylserine, phosphatidylcholine and sphingomyelin and sphingosine, all purchased from Sigma-Aldrich (Munich, Germany).

### Sphingosine and cholesterol quantitation

Preliminary results showed that the limit of quantification of sphingosine was above the typical concentration of sphingosine in erythrocyte membranes. For this reason, the levels of sphingosine were quantified in spiked (with addition of 5μg sphingosine) samples. For cholesterol there was no such limitation, and its levels were measured without spiking.

### Erythrocyte lysis

1 milliliter of packed erythrocytes was lysed with triton X-100 at a final concentration of 0.01% v/v^18^.

### Determination of MCP1 and CD47 levels

Levels of MCP1 and CD47 were determined by ELISA in erythrocyte lysates, according to the manufacturer’s instructions (OriGene, USA).

### Statistical analysis

Results were expressed as mean ± standard deviation, unless otherwise stated. Since the sample size was relatively small, a Bayesian approach for statistical analysis was employed. Bayesian analysis is much more suited to provide meaningful results for small datasets, since it does not assume large sample sizes. Thus, smaller datasets can be analyzed while retaining statistical power and precision^19,20^.

Statistical analysis was performed with the R programming language v. 4.0^21^. Testing for difference between means was performed using JAGS^22^ as implemented in the rjags package. A hierarchical model was used, assuming mean priors for the two groups that can be close or far apart. Results were reported as the probability P of the difference between the means of healthy subjects and NASH patients being less than zero, P(Healthy<NASH), or greater than zero, P(Healthy>NASH). This is more intuitive than the more convoluted meaning of the frequently used p value, which is the probability of observing the data, assuming the null hypothesis (no difference between means) is correct. Probabilities above 90% were considered statistically significant.

## RESULTS

### Characteristics of the sphingosine and cholesterol quantification method

The TLC for the quantification of sphingosine was linear through the range 1.25 – 5 μg/spot with a sensitivity of 0.90 μg/spot. The precision coefficient of variation was <2%. The limit of detection (LOD) and limit of quantification (LOQ) were 0.75 μg/spot and 1.21μg/spot, respectively.

For cholesterol, the sensitivity of the method was 0.99 μg/spot, and it was linear through the range 0.5 – 2.5 μg/spot. The precision coefficient of variation was <5%. The LOD and LOQ were 0.44 μg/spot and 4.18 μg/spot.

### Membrane CD47 levels

Erythrocyte CD47 levels were decreased in the erythrocyte membranes of NASH patients (844±409 pg/ml) compared to healthy controls (2969±1936 pg/ml) (P=99.1%) (Figure 1A and Table 2). These results indicate reduced antiphagocytic signals on erythrocytes from NASH patients.

**Table 2.** Levels (mean ± standard deviation) of measured parameters in NASH patients and healthy control subjects. The probability P is the Bayesian posterior probability that the mean of healthy group is lower (of higher for CD47) than the mean of NASH patients.

| **Parameter** | **Healthy** | **NASH** | **Probability** |
| --- | --- | --- | --- |
| CD47 | 2969±1936 | 844±409 | P(Healthy>NAFLD)=99.1% |
| MCP1 | 230±117 | 389±255 | P(Healthy<NAFLD)=88.9% |
| Sphingosine | 262±26 | 294±26 | P(Healthy<NAFLD)=93.6% |
| Cholesterol | 720±200 | 937±122 | P(Healthy<NAFLD)=95.9% |

**Figure 1.**
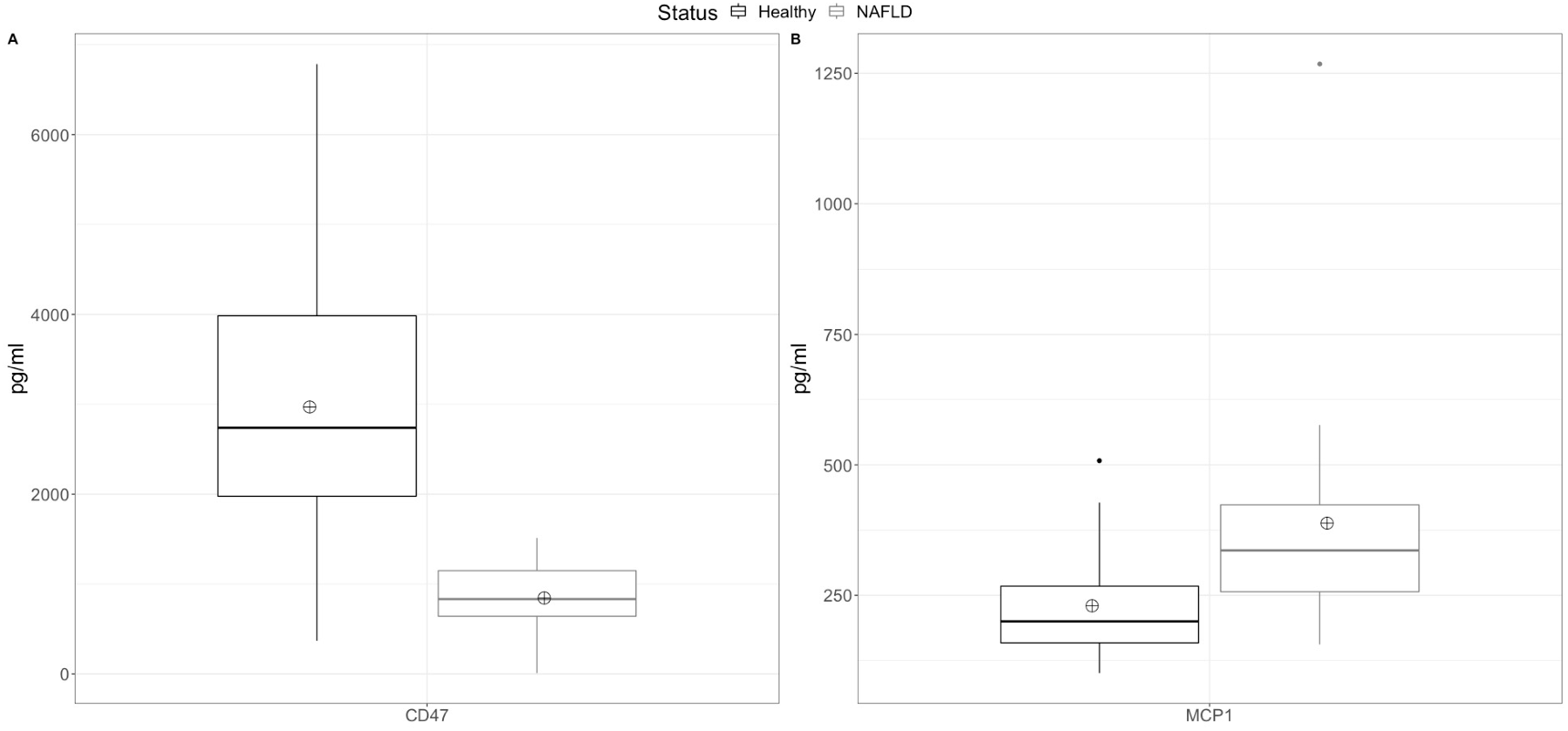
Boxplot of CD47 (A) and MCP1 (B) levels in erythrocyte membranes. The line inside the box is the median. The bottom and top edges of the box correspond to the 25^th^ and 75^th^ percentile, respectively. The box represents the interquartile range (IQR). The lower and upper whiskers are the minimum-1.5×IQ and maximum+1.5×IQR, respectively. Points outside the whiskers are outliers. The crossed circle represents the mean.

### Membrane MCP1 levels

MCP1 levels were higher in the erythrocyte membranes of patients with NASH (389±255 pg/ml), compared to healthy controls (230±117 pg/ml) (Figure 1B and Table 2). However, this difference was of marginal statistical significance (P=88.9%).

### Sphingosine levels in erythrocyte membranes and ECM

No difference in the sphingosine levels was observed between the ECM from NASH patients (4.5±1.2 μg/ml) compared to healthy controls (4.6±2.0 μg/ml). In erythrocyte membranes there was a statistically significant (P=93.6%) elevation of sphingosine in NASH patients (294±26 μg/ml packed erythrocytes), compared to healthy controls (262±26 μg/ml packed erythrocytes) (Figure 2A and Table 2).

**Figure 2.**
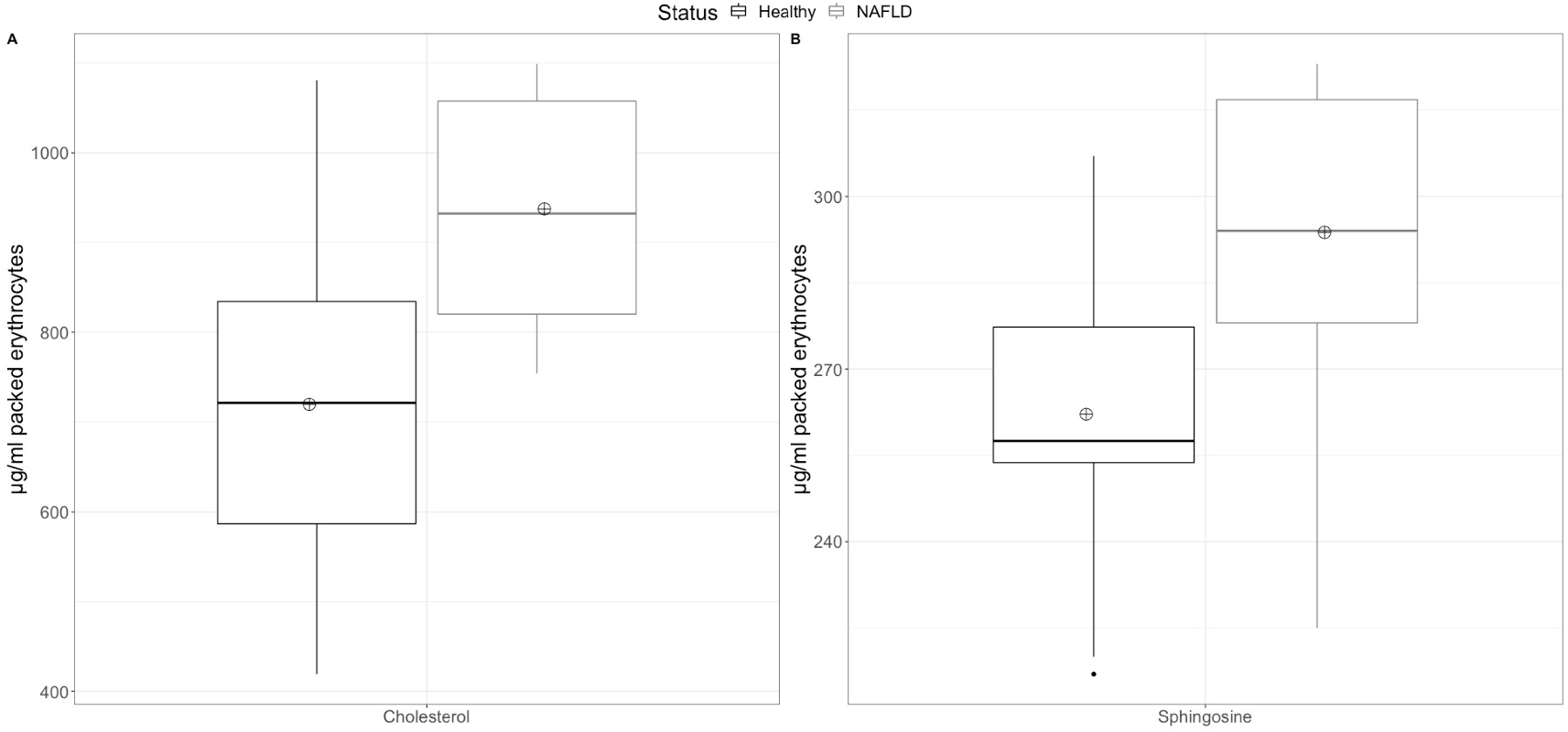
Boxplot of sphingosine (A) and cholesterol (B) levels in erythrocyte membranes. The line inside the box is the median. The bottom and top edges of the box correspond to the 25^th^ and 75^th^ percentile, respectively. The box represents the interquartile range (IQR). The lower and upper whiskers are the minimum-1.5×IQ and maximum+1.5×IQR, respectively. Points outside the whiskers are outliers. The crossed circle represents the mean.

### Sphingosine kinase inhibition

To further elaborate on the mechanism of sphingosine accumulation on erythrocytes, we decided to study the effect of sphingosine kinase inhibition on the erythrocyte sphingosine content. Inhibition of erythrocyte sphingosine kinase led to accumulation of sphingosine from 255±24 (no inhibitor present) to 375±31 μg/ml packed erythrocytes (P=99.3%).

### Membrane cholesterol levels

In erythrocyte membranes there was a statistically significant (P=95.9%) decline in cholesterol levels in healthy controls (720±200 μg/ml packed erythrocytes), compared to NASH patients (937±122 μg/ml packed erythrocytes) (Figure 2B and Table 2).

## Discussion

Erythrophagocytosis in the liver represents a highly contributing mechanism to the pathogenesis of steatohepatitis^13^. Previous research work has shown that phosphatidylserine exposure in erythrocytes and subsequent recognition by Kupffer cells, mediates this process. However, apart from the “eat me” signal (phosphatidylserine exposure), erythrophagocytosis is also controlled by a “do not eat me” signal, CD47. In addition, chemotactic molecules, like MCP1, can participate in efferocytosis, functioning as “find me” signals^23^. Indeed, increased MCP1 release from red blood cells of NASH patients^14^ has been reported. In addition to the “find me”, “eat me”, and “do not eat me” signals, phagocytized cells also release “goodbye” signals: metabolites that regulate cellular function^16^. In the context of NASH, erythrophagocytosis has been associated with enhanced hepatic inflammation and fibrosis, as a result of iron accumulation^13^. However, other metabolites that are found in red blood cells, like sphingosine and cholesterol could also be involved in the process. For example, cholesterol accumulation in Kupffer cells is implicated in enhanced inflammation^24^. For these reasons, the levels of CD47, MCP1, sphingosine and cholesterol of erythrocytes from NASH patients were measured in this study.

Reduced CD47 levels have previously been found in obese individuals^25^ and atherosclerosis patients^26^. In this study, it was shown that erythrocytes from NASH patients contain reduced CD47 levels, which could also enhance the augmented hepatic erythrophagocytosis during NASH. Reduced CD47 levels on erythrocytes could also be involved in the regulation of dendritic cell maturation^27^. The decline of the CD47 levels could be attributed to oxidative stress^26^ and erythrocyte’s TLR9 activation^28^. It is also of interest that recently, mannose was also found to represent an important signal for erythrophagocytosis^29^. This mechanism also merits further investigation in the future.

MCP1 levels in erythrocyte membranes of NASH patients were slightly increased, albeit to marginally non-statistically significant levels. However, increased membrane bound MCP1, in combination to other stimulatory factors, like ATP acting on P2X7 (unpublished results), can promote high MCP1 release from NASH patients’ erythrocytes^14^.

We next examined sphingosine levels in NASH patients and healthy controls. In NASH, there is increased serum acid sphingomyelinase^30^, which breaks sphingomyelin to phosphocholine and ceramide. Sphingomyelin hydrolysis in the erythrocyte membrane can occur as a result of inflammation-induced increased activity of plasma sphingomyelinase^31^. This event leads to the formation of ceramide^31^, sphingosine, and release of microvesicles^32^. Interestingly, erythrocyte sphingolipid metabolism can result in the formation of sphingosine 1-phosphate (S1P)^9^, a bioactive lipid mediator, implicated in the pathogenesis of NASH^33^. It has previously been shown that erythrocyte membranes of NASH patients, contain decreased levels of sphingomyelin^34^, and that erythrocyte-conditioned media do not contain elevated S1P^14^. Hence, we wondered, if there is sphingosine accumulation in the erythrocyte membrane in the context of NASH.

It was shown that sphingosine levels in the erythrocyte-conditioned media of NASH patients remain unaltered, while there are increased sphingosine levels in the erythrocyte membrane. Previous studies have shown that sphingosine accumulation in red blood cells triggers eryptosis^35^, through activation of calcium channels and subsequent activation of scramblase. Thus, eryptosis in metabolic steatohepatitis, could be also triggered by sphingosine accumulation, besides oxidative stress^13^.

Erythrocytes use the extracellular sphingosine for the synthesis of S1P^36^. Our results show that inhibition of sphingosine kinase *in vitro* can lead to sphingosine accumulation in erythrocytes. This could explain both sphingosine accumulation and the unaltered S1P release from erythrocytes of NASH patients. Nevertheless, sphingosine in the erythrocyte membrane could be utilized for the synthesis of S1P in the liver after efferocytosis^37^.

Red blood cells are also important players of the reverse cholesterol transport^9^. Since, cholesterol accumulation contributes to the inflammatory process of NASH, we wondered whether erythrocyte membranes of NASH patients contain increased cholesterol levels. A previous study showed that erythrocytes from NASH patients do not release increased cholesterol to the growth media^14^. Hence, our results here of increased membrane cholesterol could indicate that increased efferocytosis of erythrocytes in the liver of NASH patients could result in augmented cholesterol disposition in Kupffer cells.

Collectively, our results elucidate the molecular basis of erythrophagocytosis-induced hepatic inflammation in NASH (Figure 3). However, we should keep in mind that in general, exposed phosphatidylserine^38^ and erythrophagocytosis are anti-inflammatory signals^39^. The group of Angelo D’Alessandro have shown that erythrophagocytosis from macrophages leads to an alternatively activated phenotype^39^. However, it is possible that it is the cargo of phagocytized erythrocytes and the pre-conditioning of macrophages with pro-inflammatory molecules, that determine the final result of erythrophagocytosis. Hence, our results of increased sphingosine, cholesterol and MCP1 in erythrocytes of NASH patients explain why erythrophagocytosis is pro-inflammatory in the context of NASH.

**Figure 3.**
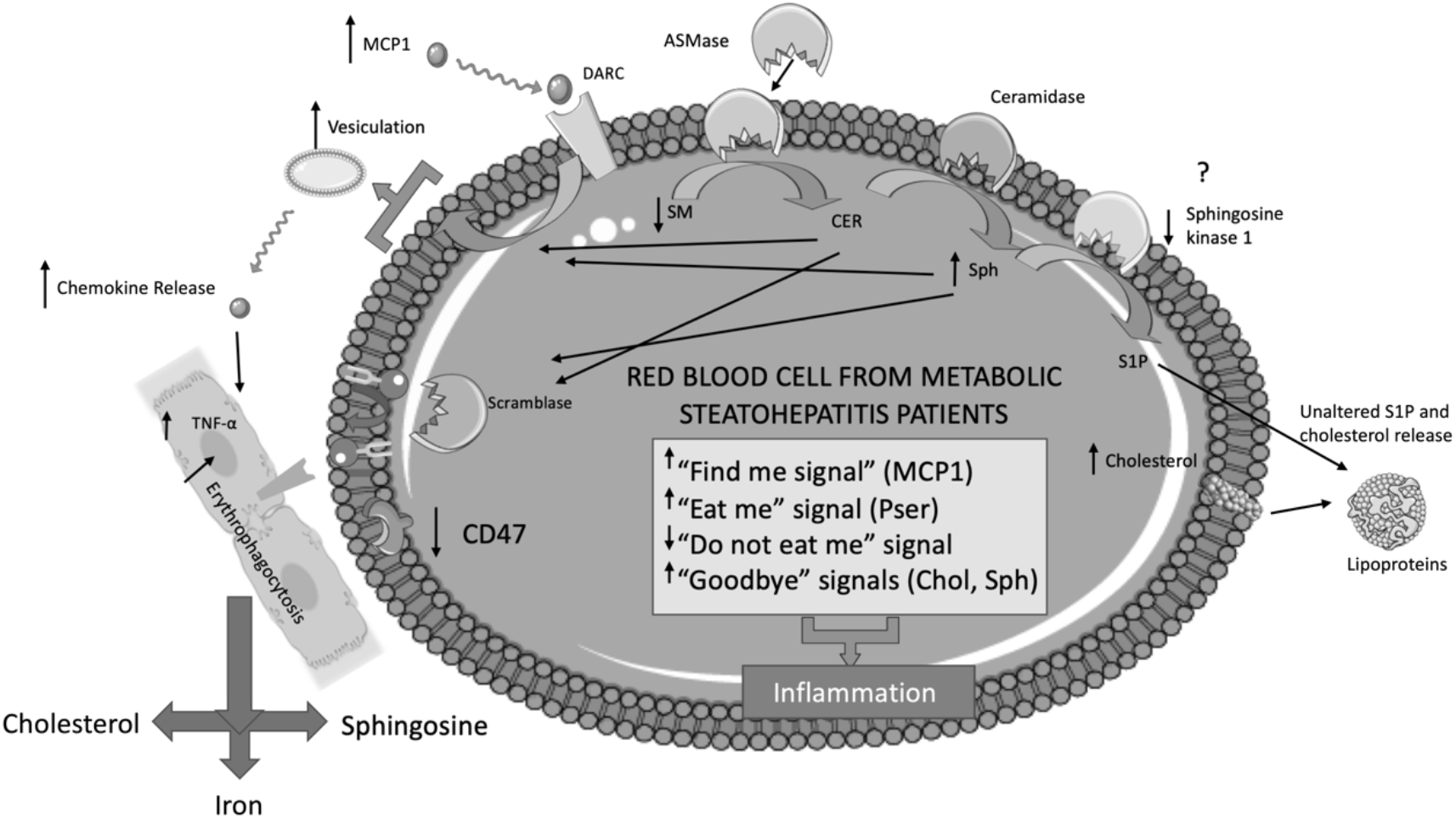
Schematic mechanism of inflammation pathways in NASH. In NASH, there is increased serum acid sphingomyelinase (ASMase)^30^, which breaks sphingomyelin (SM) to phosphocholine and ceramide (CER). This enzyme can act on the erythrocyte membrane^31^ leading to reduction of sphingomyelin^34^ and release of microvesicles^32^. Microvesicles release chemokines such as MCP1, which could activate macrophages^14^. Erythrocyte sphingolipid metabolism can result in the formation of sphingosine (Sph)^32^ and sphingosine 1-phosphate (S1P)^9^. However, it has previously been shown that erythrocyte-conditioned media do not contain elevated S1P, possibly because of inhibition of sphingosine kinase 1. Sphingosine accumulation in red blood cells triggers eryptosis^35^, through activation of calcium channels and subsequent activation of scramblase. Phosphatidylserine exposure^13^ along with reduced membrane CD47 levels can promote erythrophagocytosis in the liver. Once erythrophagocytosis has occurred, increased membrane cholesterol and sphingosine, along with iron disposition could augment inflammation. This figure was created using Servier Medical Art templates, which are licensed under a Creative Commons Attribution 3.0 Unported License; https://smart.servier.com.

## Conclusion

Our results imply that erythrocytes of NASH patients release chemotactic “find me” signals (MCP1), while containing reduced “do not eat me” signals (CD47). These molecules can lead to erythrophagocytosis. Next, increased “goodbye” signals (sphingosine and cholesterol) could augment inflammation. Further research on the implicated molecular mechanisms of these immunometabolic interactions, could lead to novel therapeutic targets for NASH.

## Data Availability

All data produced in the present work are contained in the manuscript

